# The prognostic value of transthoracic echocardiography findings in hospitalized adult patients with COVID-19: A single-center retrospective analysis

**DOI:** 10.1101/2022.08.07.22278506

**Authors:** Hary Sakti Muliawan, Raksheeth Agarwal, Raka Aldy Nugraha, Gatut Priyonugroho, Siti Hertine, Sony Hilal Wicaksono, Prima Almazini, Dian Zamroni

## Abstract

**Background:** Cardiac involvement in coronavirus disease 2019 (COVID-19) is associated with poor outcomes. Transthoracic echocardiography (TTE) can be used to assess cardiac structure and function non-invasively, and has been shown to influence management in COVID-19.

**Objectives:** We aim to investigate the prognostic value of TTE findings in hospitalized adults with confirmed COVID-19.

**Methods:** All consecutive hospitalized adult patients with confirmed COVID-19 who underwent TTE assessment between 3rd April 2020 – 6th April 2021 were included. Comprehensive clinical data including TTE findings were collected from electronic medical records. Patients with mild-moderate and severe-critical COVID-19 were compared. Within the severe-critical group, patients who survived hospitalization and died were compared. Further analyses were conducted after matching for age >60 years, obesity, and diabetes.

**Results:** A total of 488 COVID-19 patients were included in this study; 202 with mild-moderate and 286 severe-critical disease. All mild-moderate patients and 152 severe-critical patients survived hospitalization. In the matched cohorts, TTE findings associated with severe-critical COVID-19 included left ventricular (LV) hypertrophy (OR: 1.91; CI: 1.21 – 3.02), LV diastolic dysfunction (OR: 1.55; CI: 1.00 – 2.38), right ventricular (RV) dysfunction (OR: 3.86; CI: 1.06 – 14.08), wall motion abnormalities (WMAs) (OR: 2.76; CI: 1.28 – 5.96), and any TTE abnormalities (OR: 2.99; CI: 1.73 – 5.17). TTE findings associated with mortality included RV dysfunction (OR: 3.53; CI: 1.12 – 11.19) and WMAs (OR: 2.63; CI: 1.26 – 5.49).

**Conclusion:** TTE is a non-invasive modality that can potentially be used for risk-stratification of hospitalized COVID-19 patients. These findings must be confirmed in larger prospective studies.

## 1. Introduction

The coronavirus disease 2019 (COVID-19) caused by severe acute respiratory syndrome coronavirus 2 (SARS-CoV-2) was first described in December 2019. The disease has since spread rapidly, being declared a pandemic in March 2020 which is currently ongoing.^1^ More than 580 million cases and 6.4 million deaths have been reported worldwide.^2,3^ The clinical spectrum of COVID-19 varies widely, from asymptomatic infection to critically severe disease characterized by respiratory failure, septic shock, and multi-organ dysfunction.^4^ Rapid community transmission of COVID-19 can overwhelm healthcare systems and cause crises, especially in low-resource settings.^5,6^ Hence, prognostic markers that can predict the clinical trajectory of patients are valuable, as they allow for risk-stratification and efficient resource allocation.

COVID-19 is primarily a respiratory condition, but has significant effects on multiple organ systems. Cardiovascular complications of COVID-19 include myocardial injury, acute heart failure, and acute coronary syndrome, and their incidence is associated with a poor prognosis. Besides this, the presence of pre-existing cardiovascular comorbidities also predicts worse clinical outcomes.^7–9^ Therefore, identification of pre-existing and *de novo* cardiovascular abnormalities could provide valuable prognostic information in COVID-19 patients. Transthoracic echocardiography (TTE) is a quick, non-invasive, and widely available tool for the assessment of cardiac structure and function. In a previous global study on 1216 patients with COVID-19, TTE findings changed management in 33% of the patients, demonstrating the value of this modality.^10^ In this retrospective study, we aim to investigate the prognostic value of TTE findings in hospitalized adults with confirmed COVID-19.

## 2. Materials and Methods

### 2.1 Study population and design

This single-center retrospective observational study was conducted at Universitas Indonesia Hospital, a designated COVID-19 referral hospital in Indonesia. All consecutive hospitalized adult patients with confirmed COVID-19 who underwent TTE assessment between 3^rd^ April 2020 – 6^th^ April 2021 were included in this study. A confirmed case of COVID-19 was defined by a positive reverse-transcriptase polymerase chain reaction (RT-PCR) test for SARS-CoV-2. Patients under the age of 18 or with negative RT-PCR results were excluded. This retrospective study was performed in line with the principles of the Declaration of Helsinki, and ethical approval was granted by the Institutional Review Board of Universitas Indonesia Hospital (Ref: 2022-01-135).

### 2.2 Transthoracic echocardiography

Patients with appropriate indications underwent focused TTE assessments during hospitalization. The decision for whether a TTE study is needed was made by a clinical cardiologist as a part of the multidisciplinary team. All TTE assessments were conducted by cardiologists experienced in this technique. According to recommendations from the American Society of Echocardiography (ASE) and the Indonesian Heart Association, TTE assessments were focused as necessary to aid management decisions.^11,12^ Other appropriate infection control and protective precautions were taken as recommended by the ASE.^11^

All TTE findings were recorded on a standardized echocardiography reporting form. The following TTE parameters were extracted for this study: left atrial (LA) dilatation, left ventricle (LV) hypertrophy, LV ejection fraction, LV systolic function, right ventricle (RV) function, wall motion abnormalities (WMAs), and LV diastolic function. LA dilation was defined as an anteroposterior LA diameter > 40mm measured in the parasternal long-axis view, LA volume index >34mL/m^2^, or by visual estimation. LV measurements were made in the parasternal long-axis view; LV hypertrophy (LVH) was defined as an LV mass > 95 g/m^2^ for women, and >115 g/m^2^ for men. LV ejection fraction (LVEF) was measured by Simpson’s biplane method, TEICHOLZ quantification method on M-Mode, or estimated visually when image quality did not allow for accurate quantification. LV systolic dysfunction was defined as an LVEF < 50%. RV dysfunction was defined by an abnormal tricuspid annular plane systolic excursion (TAPSE) (<17mm) or RV dilation (RV basal diameter >41mm or visual estimation). LV diastolic dysfunction was defined according to the structural and functional criteria set by the ASE.^13^ Any WMAs were also recorded.

### 2.3 Data collection

All data was extracted from the hospital’s secure electronic medical records (EMR) system. The extracted data included demographic information, peak severity of COVID-19 during hospitalization, final outcome of disease (survival or death), and presence of comorbidities including obesity, hypertension, diabetes, asthma, chronic obstructive pulmonary disease (COPD), chronic kidney disease (CKD), hypertensive heart disease (HHD), congestive heart failure (CHF), coronary artery disease (CAD), and malignancy. Severity of COVID-19 was graded as mild, moderate, severe, or critical, as defined by the National Institutes of Health practice guidelines.^4^ TTE parameters were extracted from the patients’ standardized echocardiography report forms, also available on the EMR system.

### 2.4 Clinical outcomes and statistical analysis

Comparisons were made between patients with mild-moderate and severe-critical disease. Within the severe-critical group, further comparisons were made between patients who died and patients who survived hospitalization. Further analyses were conducted after matching for age >60 years, obesity, and diabetes.

Normality of continuous variables was assessed using the Shapiro-Wilk test. Continuous variables are presented as mean (95% confidence interval) when normally distributed, or median (interquartile range) when non-normally distributed. Categorical variables are expressed as counts and percentages. Continuous variables were compared using the Student’s T-test when normally distributed, or the Mann-Whitney’s U-test when non-normally distributed. Categorical variables were compared using the chi-square test or the Fischer’s exact test. Odds ratios were calculated for all echocardiography parameters. A p-value of <0.05 was considered statistically significant. All statistical analyses were conducted using IBM SPSS Statistics 26 (IBM Corporations, USA).

## 3. Results

### 3.1 Study population

A total of 488 hospitalized adult patients with confirmed COVID-19 underwent focused TTE assessments between 3^rd^ April 2020 and 6^th^ April 2021 (Figure 1). All patients with mild-moderate disease survived hospitalization. The most common indications for TTE were a history or suspicion of CHF (N=172) or HHD (N=164). Other indications included a history of CAD (N=59), diabetes (N=15), new onset dyspnea (N=10), or obesity (N=2). The TTE indication for 66 patients was undocumented.

**Fig. 1.**
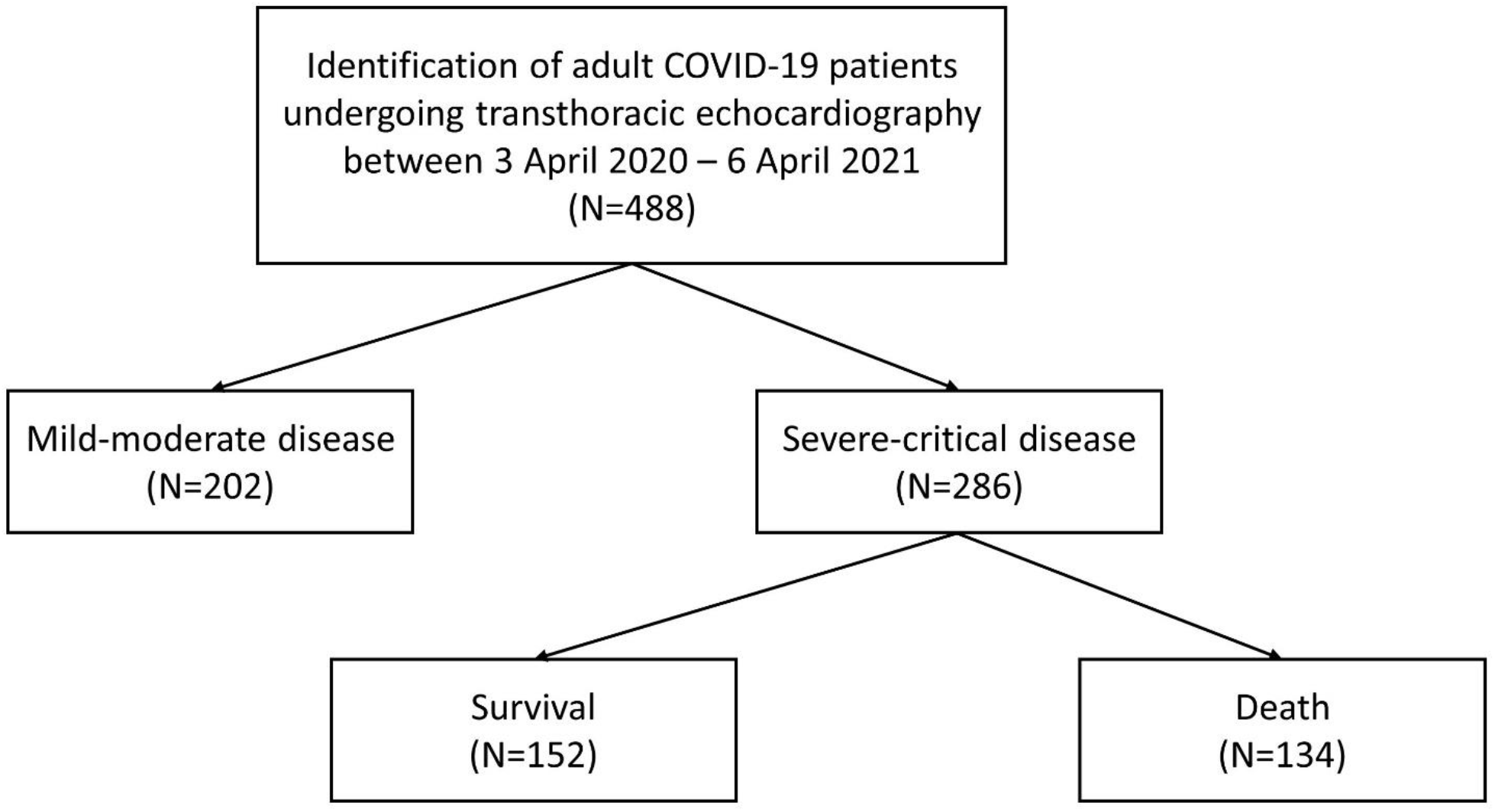
Study population

### 3.2 Severity analysis

During hospitalization 202 patients experienced mild-moderate disease, and 286 patients experienced severe-critical disease. The demographic characteristics, comorbidities, and echocardiographic findings of these patients are presented in table 1. Patients with severe-critical disease were older and had a higher frequency of obesity, diabetes, CKD, and CHF. TTE assessments showed that this group had higher odds of experiencing any abnormalities, LVH, RV dysfunction, WMAs, and LV diastolic dysfunction

**Table 1.**
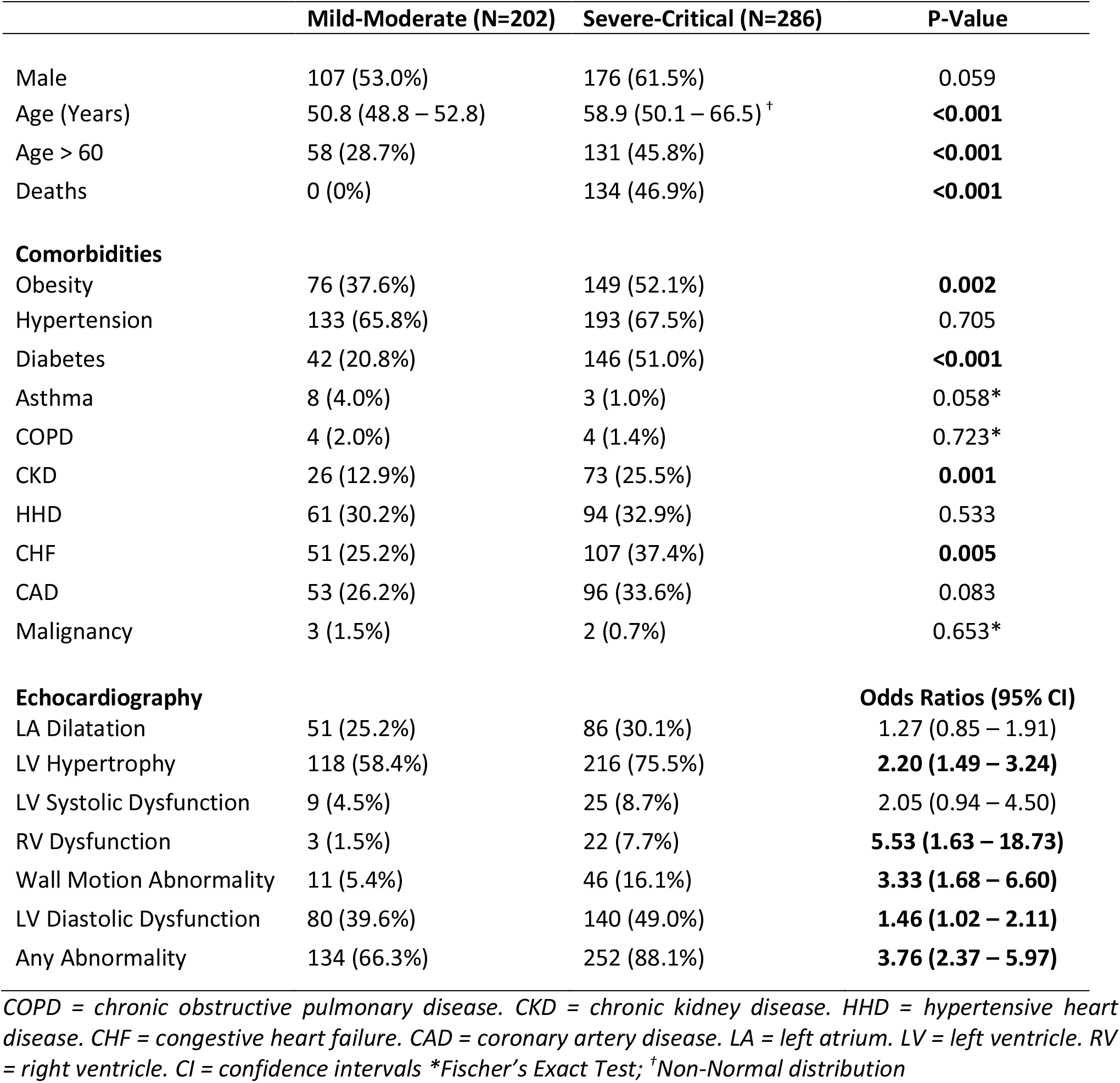
Severity analysis for the entire population.

|  | **Mild-Moderate (N=202)** | **Severe-Critical (N=286)** | **P-Value** |
| --- | --- | --- | --- |
| Male | 107 (53.0%) | 176 (61.5%) | 0.059 |
| Age (Years) | 50.8 (48.8 – 52.8) | 58.9 (50.1 – 66.5) <sup>†</sup> | **<0.001** |
| Age > 60 | 58 (28.7%) | 131 (45.8%) | **<0.001** |
| Deaths | 0 (0%) | 134 (46.9%) | **<0.001** |
| **Comorbidities** |  |  |  |
| Obesity | 76 (37.6%) | 149 (52.1%) | **0.002** |
| Hypertension | 133 (65.8%) | 193 (67.5%) | 0.705 |
| Diabetes | 42 (20.8%) | 146 (51.0%) | **<0.001** |
| Asthma | 8 (4.0%) | 3 (1.0%) | 0.058* |
| COPD | 4 (2.0%) | 4 (1.4%) | 0.723* |
| CKD | 26 (12.9%) | 73 (25.5%) | **0.001** |
| HHD | 61 (30.2%) | 94 (32.9%) | 0.533 |
| CHF | 51 (25.2%) | 107 (37.4%) | **0.005** |
| CAD | 53 (26.2%) | 96 (33.6%) | 0.083 |
| Malignancy | 3 (1.5%) | 2 (0.7%) | 0.653* |
| **Echocardiography** |  |  | **Odds Ratios (95% CI)** |
| LA Dilatation | 51 (25.2%) | 86 (30.1%) | 1.27 (0.85 – 1.91) |
| LV Hypertrophy | 118 (58.4%) | 216 (75.5%) | **2.20 (1.49 – 3.24)** |
| LV Systolic Dysfunction | 9 (4.5%) | 25 (8.7%) | 2.05 (0.94 – 4.50) |
| RV Dysfunction | 3 (1.5%) | 22 (7.7%) | **5.53 (1.63 – 18.73)** |
| Wall Motion Abnormality | 11 (5.4%) | 46 (16.1%) | **3.33 (1.68 – 6.60)** |
| LV Diastolic Dysfunction | 80 (39.6%) | 140 (49.0%) | **1.46 (1.02 – 2.11)** |
| Any Abnormality | 134 (66.3%) | 252 (88.1%) | **3.76 (2.37 – 5.97)** |

*COPD = chronic obstructive pulmonary disease. CKD = chronic kidney disease. HHD = hypertensive heart disease. CHF = congestive heart failure. CAD = coronary artery disease. LA = left atrium. LV = left ventricle. RV = right ventricle. CI = confidence intervals \*Fischer's Exact Test; <sup>†</sup>Non-Normal distribution*

After matching for old age (>60 years), obesity, and diabetes (table 2), CHF was the only comorbidity with a significant difference between the two groups. The severe-critical group continued to have higher odds of any abnormality on TTE, and higher odds of LVH, RV dysfunction, WMAs, and LV diastolic dysfunction.

**Table 2.** Severity analysis for matched cohort.

|  | **Mild-Moderate (N=167)** | **Severe-Critical (N=167)** | **P-Value** |
| --- | --- | --- | --- |
| Male | 89 (53.3%) | 104 (62.3%) | 0.097 |
| Age (Years) | 52.6 (50.3 – 54.9) | 55.1 (53.2 – 57.0) | 0.101 |
| Age > 60 | 58 (34.7%) | 58 (34.7%) | - |
| Deaths | 0 (0%) | 47 (28.1%) | **<0.001** |
| **Comorbidities** |  |  |  |
| Obesity | 72 (43.1%) | 72 (43.1%) | - |
| Hypertension | 113 (67.7%) | 110 (65.9%) | 0.727 |
| Diabetes | 42 (25.1%) | 42 (25.1%) | - |
| Asthma | 7 (4.2%) | 2 (1.2%) | 0.174* |
| COPD | 4 (2.4%) | 2 (1.2%) | 0.685* |
| CKD | 22 (13.2%) | 34 (20.4%) | 0.079 |
| HHD | 55 (32.9%) | 57 (34.1%) | 0.817 |
| CHF | 39 (23.4%) | 65 (38.9%) | **0.002** |
| CAD | 44 (26.3%) | 55 (32.9%) | 0.188 |
| Malignancy | 3 (1.8%) | 2 (1.2%) | 1.000 |
| **Echocardiography** |  |  | **Odds Ratios (95% CI)** |
| LA Dilatation | 39 (23.4%) | 52 (31.1%) | 1.48 (0.91 – 2.41) |
| LV Hypertrophy | 98 (58.7%) | 122 (73.1%) | **1.91 (1.21 – 3.02)** |
| LV Systolic Dysfunction | 8 (4.8%) | 12 (7.2%) | 1.54 (0.61 – 3.87) |
| RV Dysfunction | 3 (1.8%) | 11 (6.6%) | **3.86 (1.06 – 14.08)** |
| Wall Motion Abnormality | 10 (6.0%) | 25 (15.0%) | **2.76 (1.28 – 5.96)** |
| LV Diastolic Dysfunction | 69 (41.3%) | 87 (52.1%) | **1.55 (1.00 – 2.38)** |
| Any Abnormality | 113 (67.7%) | 144 (86.2%) | **2.99 (1.73 – 5.17)** |

*COPD = chronic obstructive pulmonary disease. CKD = chronic kidney disease. HHD = hypertensive heart disease. CHF = congestive heart failure. CAD = coronary artery disease. LA = left atrium. LV = left ventricle. RV = right ventricle. CI = confidence intervals \*Fischer's Exact Test*

### 3.3 Mortality analysis

Of the 286 patients with severe-critical disease, 152 survived hospitalization and 134 died. Their demographic characteristics, comorbidities, and echocardiographic findings are given in table 3. Patients who died had a higher frequency of diabetes, CKD, and CAD. On TTE, this group had higher odds of RV dysfunction and WMAs.

**Table 3.** Mortality analysis for all severe-critical patients.

|  | **Survival (N=152)** | **Death (N=134)** | **P-Value** |
| --- | --- | --- | --- |
| Male | 92 (60.5%) | 84 (62.7%) | 0.708 |
| Age (Years) | 55.8 (53.8 – 57.7) | 59.7 (57.8 – 61.7) | **0.005** |
| Age > 60 | 64 (42.1%) | 67 (50.0%) | 0.181 |
| **Comorbidities** |  |  |  |
| Obesity | 73 (48.0%) | 76 (56.7%) | 0.142 |
| Hypertension | 105 (69.1%) | 88 (65.7%) | 0.539 |
| Diabetes | 68 (44.7%) | 78 (58.2%) | **0.023** |
| Asthma | 1 (0.7%) | 2 (1.5%) | 0.601* |
| COPD | 2 (1.3%) | 2 (1.5%) | 1.000* |
| CKD | 23 (15.1%) | 50 (37.3%) | **<0.001** |
| HHD | 51 (33.6%) | 43 (32.1%) | 0.793 |
| CHF | 61 (40.1%) | 46 (34.3%) | 0.312 |
| CAD | 43 (28.3%) | 53 (39.6%) | **0.044** |
| Malignancy | 1 (0.7%) | 1 (0.7%) | 1.000* |
|  |  |  | **Odds Ratios (95% CI)** |
| **Echocardiography** |  |  |  |
| LA Dilatation | 42 (27.6%) | 44 (32.8%) | 1.28 (0.77 – 2.12) |
| LV Hypertrophy | 115 (75.7%) | 101 (75.4%) | 0.99 (0.57 – 1.69) |
| LV Systolic Dysfunction | 9 (5.9%) | 16 (11.9%) | 2.15 (0.92 – 5.05) |
| RV Dysfunction | 7 (4.6%) | 15 (11.2%) | **2.61 (1.03 – 6.61)** |
| Wall Motion Abnormality | 13 (8.6%) | 33 (24.6%) | **3.49 (1.75 – 6.97)** |
| LV Diastolic Dysfunction | 76 (50.0%) | 64 (47.8%) | 0.91 (0.57 – 1.46) |
| Any Abnormality | 130 (85.5%) | 122 (91.0%) | 1.72 (0.82 – 3.63) |

*COPD = chronic obstructive pulmonary disease. CKD = chronic kidney disease. HHD = hypertensive heart disease. CHF = congestive heart failure. CAD = coronary artery disease. LA = left atrium. LV = left ventricle. RV = right ventricle. CI = confidence intervals \*Fischer's Exact Test*

After matching for age, obesity, and diabetes (table 4), CKD was the only significantly different comorbidity between the survival and death groups. Odds ratios for RV dysfunction and WMAs on TTE remained statistically significant.

**Table 4.** Mortality analysis for matched severe-critical patients.

|  | **Survived (N=116)** | **Death (N=116)** | **P-Value** |
| --- | --- | --- | --- |
| Male | 69 (59.5%) | 77 (66.4%) | 0.277 |
| Age (Years) | 56.3 (54.2 – 58.4) | 58.8 (56.7 – 60.9) | 0.094 |
| Age > 60 | 51 (44.0%) | 51 (44.0%) | - |
| **Comorbidities** |  |  |  |
| Obesity | 60 (51.7%) | 60 (51.7%) | - |
| Hypertension | 78 (67.2%) | 76 (65.5%) | 0.781 |
| Diabetes | 60 (51.7%) | 60 (51.7%) | - |
| Asthma | 1 (0.9%) | 2 (1.7%) | 1.000* |
| COPD | 1 (0.9%) | 1 (0.9%) | 1.000* |
| CKD | 21 (18.1%) | 42 (36.2%) | **0.002** |
| HHD | 41 (35.3%) | 39 (33.6%) | 0.782 |
| CHF | 51 (44.0%) | 41 (35.3%) | 0.180 |
| CAD | 35 (30.2%) | 49 (42.2%) | 0.056 |
| Malignancy | 1 (0.9%) | 1 (0.9%) | 1.000* |
|  |  |  | **Odds Ratios (95% CI)** |
| **Echocardiography** |  |  |  |
| LA Dilatation | 33 (28.4%) | 34 (29.3%) | 1.04 (0.59 – 1.84) |
| LV Hypertrophy | 92 (79.3%) | 85 (73.3%) | 0.72 (0.39 – 1.32) |
| LV Systolic Dysfunction | 8 (6.9%) | 12 (10.3%) | 1.56 (0.61 – 3.97) |
| RV Dysfunction | 4 (3.4%) | 13 (11.2%) | **3.53 (1.12 – 11.19)** |
| Wall Motion Abnormality | 12 (10.3%) | 27 (23.3%) | **2.63 (1.26 – 5.49)** |
| LV Diastolic Dysfunction | 60 (51.7%) | 55 (47.4%) | 0.84 (0.50 – 1.41) |
| Any Abnormality | 101 (87.1%) | 104 (89.7%) | 1.29 (0.57 – 2.89) |

*COPD = chronic obstructive pulmonary disease. CKD = chronic kidney disease. HHD = hypertensive heart disease. CHF = congestive heart failure. CAD = coronary artery disease. LA = left atrium. LV = left ventricle. RV = right ventricle. CI = confidence intervals \*Fischer's Exact Test*

## 4. Discussion

In this report, we describe the range of TTE abnormalities and their associations with severity and mortality in hospitalized adults with COVID-19. Old age (>60 years), obesity, and diabetes are well-known risk factors for severe COVID-19,^14,15^ which is consistent with our findings (table 1). After matching for these factors, a history of CHF and CKD were associated with severity and mortality, respectively, corroborating previous reports.^16,17^ As for TTE findings, RV dysfunction and WMAs were associated with both severity of disease and mortality. Additionally, LVH, LV diastolic dysfunction, and the presence of any abnormality were associated with severe-critical COVID-19.

Emerging evidence suggests that RV dysfunction is an important prognostic factor in COVID-19. A previous meta-analysis^18^ estimated the prevalence of RV dysfunction in COVID-19 to be 20.4% with significant variability between studies. However, most included studies were conducted on severely ill patients in the intensive care unit, which could have overestimated its true prevalence. The prevalence of RV dysfunction in our study was only 5.1%, possibly because we included all hospitalized patients, including mild cases.

In our study, RV dysfunction was associated with both severity and mortality in COVID-19, which is consistent with previous reports.^18,19^ Several pathophysiological mechanisms for the association between RV dysfunction and poor outcomes in COVID-19 have been described. Acute *cor pulmonale* is frequently observed in acute respiratory distress syndrome (ARDS), which is a key feature of critical COVID-19. This is thought to be due to the effects of hyperinflammation, cytokine storm, hypercapnia, and ventilatory strategies (e.g. driving pressure and hyperinflation) on the pulmonary vascular bed, increasing vascular resistance.^20^ Besides this, COVID-19 is associated with endothelial dysfunction, resulting in a hypercoagulable and vasoconstrictor phenotype.^21,22^ This hypercoagulable and hyperinflammatory state predisposes patients to pulmonary emboli and microthrombi, which are frequently observed in COVID-19.^23,24^ Together, these factors increase pulmonary vascular resistance and right ventricular afterload, increasing the risk of RV dysfunction. The susceptibility of the overloaded right ventricle is further amplified by direct myocardial injury occurring from various mechanisms.^24^

Besides RV dysfunction, WMAs were associated with both severity and mortality in our study. In a previous study, WMAs were observed in 15% of hospitalized adults with COVID-19, which is comparable to the prevalence in our cohort. In this previous study, some patients had new or worsened WMAs during the COVID-19 episode, while others had chronic abnormalities.^25^ WMAs in COVID-19 occur due to various etiologies, including coronary artery disease, acute myocarditis, stress (Takotsubo) cardiomyopathy, and CHF (chronic or acute).^26^ As such, the presence of WMAs in COVID-19 represents a mix of acute *de novo* and chronic cardiovascular abnormalities, both of which are associated with poor outcomes.^7,9^ Several underlying pathophysiological mechanisms for acute cardiovascular injury have been proposed, including cytokine storm, hyperinflammation, hypercoagulability, direct cellular injury, emotional stress, microvascular/ endothelial dysfunction, hypoxia, and sepsis.^26^

Our study demonstrated that LV diastolic dysfunction was associated with severe-critical disease. Previously, Stöbe, el al. found that patients requiring mechanical ventilation had a higher E/e’ ratio compared to those who did not.^27^ Furthermore, Szekely, et al. reported an association between an elevated E/e’ ratio and mortality, and a positive correlation between the E/e’ ratio and troponin levels in their COVID-19 cohort.^28^ In another study on elderly patients with echocardiography done several months (median 42.2 months) before the onset of COVID-19, previous evidence of grade 2 or 3 diastolic dysfunction was predictive of hospitalization or ARDS during the COVID-19 episode.^29^ Together, these studies suggest that diastolic dysfunction in COVID-19 may represent either acute myocardial injury or chronic comorbidity, and is associated with poor outcomes.

While current evidence on the prognostic value of LVH in COVID-19 is scarce, our study found that this echocardiographic parameter was associated with severe-critical disease. A previous study reported that LVH detected by electrocardiography (ECG) at admission was associated with a composite of all-cause mortality and/or respiratory failure during hospitalization.^30^ Ghany, et al. demonstrated that evidence of moderate or severe LVH on TTE performed several months before the onset of COVID-19 was associated with a higher risk of hospitalization and ARDS during the COVID-19 episode.^29^

LVH represents chronic adaptive hyperplasia of cardiomyocytes in response to excess pressure (e.g. hypertension, aortic stenosis) or volume overload (e.g. mitral regurgitation, aortic regurgitation), resulting in concentric or eccentric hypertrophy, respectively. Concentric hypertrophy due to chronic pressure overload often leads to diastolic dysfunction.^31^ The prevalence of chronic hypertension in our study was high (66.8%). According to the *Indonesia Family Life Survey (IFLS-5)* conducted in 2015, only 11.5% of Indonesian hypertensive patients routinely consumed antihypertensive medication, and only 14.3% had an adequately controlled blood pressure (systolic blood pressure <140 mmHg and diastolic blood pressure <90 mmHg).^32^ The prevalence of both LVH (68.4%) and LV diastolic dysfunction (45.1%) in our study were higher than previously reported in COVID-19.^28,30^ It is possible that the high prevalence of LVH and LV diastolic dysfunction represents poorly controlled hypertension in our population.

Several limitations of our study warrant a discussion. Similar to previous studies,^33,34^ the TTE performed in our patients was focused and not always comprehensive in order to reduce the risk of infection. Secondly, this study was retrospective in nature, which results in a higher risk of bias and reduced control on data availability. Thirdly, the timing of TTE in our study was variable, and hence there could be variability in disease progression between patients at the time of TTE. Fourthly, we did not differentiate patients according to the variant of SARS-CoV-2, and so the effect of different variants on TTE findings and clinical outcomes is unknown. Finally, TTE was only performed once, and no measurements were taken before or after the study. Hence, it is unknown whether the abnormalities observed were chronic or acute, or whether they persisted post-recovery. Nevertheless, the aim of our study was to investigate the prognostic value of TTE findings during hospitalization regardless of whether they were chronic or acute.

Despite these limitations, this is one of the largest real-world studies investigating the prognostic value of TTE in COVID-19,^35^ and hence provides some important insights. If feasible and safe, TTE must be conducted in hospitalized COVID-19 patients with appropriate indications, as this non-invasive modality can provide valuable prognostic information. Several studies have reported RV dysfunction and WMAs to be associated with a poor prognosis, which is consistent with our findings. Additionally, the presence of any TTE abnormality was associated with severe-critical disease, and hence can alert physicians of possible deterioration in patients. Future prospective studies investigating the prognostic value of TTE in COVID-19 must standardize the timing of measurements.

## 5. Conclusion

This retrospective observational study demonstrates that TTE abnormalities including RV dysfunction, WMAs, LVH, and LV diastolic dysfunction are associated with poor outcomes in COVID-19. TTE can potentially be used as a non-invasive modality for risk-stratification of hospitalized COVID-19 patients. These findings must be confirmed and further elaborated in future prospective studies.

## Data Availability

The data that support the findings of this study are available upon reasonable request from the corresponding author. The data are not publicly available due to privacy or ethical restrictions.

## Ethics approval

This retrospective observational study was performed in line with the principles of the Declaration of Helsinki, and ethical approval was granted by the Institutional Review Board of Universitas Indonesia Hospital (Ref: 2022-01-135).

## Author Contributions

All authors contributed to the study conception and design. Material preparation, data collection and analysis were performed by Hary Sakti Muliawan and Raksheeth Agarwal. The first draft of the manuscript was written by Raksheeth Agarwal and all authors commented on previous versions of the manuscript and revised it critically for intellectual content. All authors approved of the final manuscript to be published and agree to be accountable for all aspects of the work.

